# Task performance to discriminate mild cognitive impairment from healthy aging

**DOI:** 10.1101/2023.09.27.23296235

**Authors:** Melissa D. Stockbridge, Argye E. Hillis

**Affiliations:** Department of Neurology, Johns Hopkins University School of Medicine, Baltimore, MD 21287; Department of Physical Medicine and Rehabilitation, Johns Hopkins University School of Medicine, Baltimore, MD 21287; Department of Cognitive Science, Krieger School of Arts and Sciences, Johns Hopkins University, Baltimore, MD 21218

**Keywords:** cognitive impairment, diagnosis, evaluation, language, cognition

## Abstract

Mild cognitive impairment (MCI) is clinical diagnosis that refers to individuals whose performance is below average on standardized cognitive tests, but who otherwise function independently in instrumental activities of daily living. Few prior studies have addressed the problem of selecting the optimal combination of behavioral instruments and cutoff scores for detecting MCI in an outpatient setting. The aim of this work is to provide insight into two related questions: (1) What is the relative sensitivity and specificity of a battery of standardized tests frequently used to assess for MCI, as defined using receiver operating characteristic (ROC)-based analysis? (2) What are the optimal “cut point” scores for distinguishing patients’ mildly impaired performance based on these instruments?

Two hundred forty outpatient behavioral neurology evaluations were retrospectively analyzed. All work was conducted with the formal approval of the Johns Hopkins University School of Medicine Institutional Review Board. All instruments that were evaluated performed very well in the detection of dementia (mean AUC = 0.8). However, fewer tasks performed acceptably in the detection of MCI (mean AUC = 0.7). Instruments that performed best in the detection of MCI tended to have higher total possible scores or not to reflect a score out of a total number possible.

Cognitive screening tools, like the MMSE, did not perform well in the detection of MCI, raising important considerations for their interpretation. No one task in isolation is sufficient to detect MCI, and behavioral performance is not the only relevant consideration in differential diagnosis. However, these findings highlight the relative weakness of many assessments when used to build a comprehensive profile of a very large portion of outpatients presenting at clinic, those whose deficits are more subtle.

## Introduction

Mild cognitive impairment (MCI) is clinical diagnosis that refers to individuals whose performance on standardized cognitive tests has fallen outside of the average performance for individuals like them. People with MCI otherwise function independently in instrumental activities of daily living and do not meet criteria for a diagnosis of dementia.^1^ In practice, this frequently means task performance by people with MCI is poorer than one standard deviation below the normative performance available that permits the closest comparison to their baseline.^2^ Normative performance generally is parsed by decade of age, sometimes sex, and rarely education. The potential lack of specificity or congruency between individual characteristics influencing performance on cognitive and cognitive communication tasks in a given task or battery of tasks and available norms is well-known. It is a widely discussed problem in psychometrics that can lead to both over- and under-identification of clinical profiles, such as MCI.^3-5^

Concurrently, the identification and understanding of MCI faces another set of challenges related to its nature and the limitations of tools used to detect it. Because its definition is linked inextricably to performance, it is inherently heterogeneous in underlying etiology. People can have mild cognitive impairment in the absence of any observable clinical change on standard magnetic resonance neuroimaging. Due to the heterogeneous etiology, some individuals with MCI improve over time back to performance within normal limits (WNL) for their age, while some appear to remain statically impaired, and some progress further from normative performance to dementia. The ability to distinguish among these fates is a high-stakes, high-investment line of research in the clinical cognitive and neuropsychological sciences.^6-8^

In the meantime, as these predictive models continue to be refined, the identification of MCI faces another problem. That is, individuals who report cognitive change in an outpatient setting first are evaluated in most cases using tests designed for the detection of *dementia*, not the detection of MCI. While it may appear at first glance to be a minor point of concern, this can be conceptualized as another kind of normative performance mismatch to the individual, akin to poor normative comparators for an individual’s education or culture. Tests designed for the quantification of larger changes do not necessarily perform well when detecting small changes, especially if the individual is non-central to the assessment’s normative sample in other ways, for example, being of a younger age than most of the sample. In these circumstances, “normal” performance can equate to ceiling performance (see examples in Table 1), rendering it difficult if not impossible to verify whether an individual’s subjective impression of cognitive change is corroborated by changes in performance (either at the first evaluation or subsequent visits). Few prior studies have addressed the problem of selecting the optimal combination of behavioral instruments and cutoff scores for distinguishing typical and atypical performance associated with MCI across domains in a typical outpatient setting. This gap in the literature leads to the use of extended batteries that often are impractical to implement and challenging for patients, particularly those further in their disease progression, to complete. The aim of this work is to provide insight into two related questions:

**Table 1:**
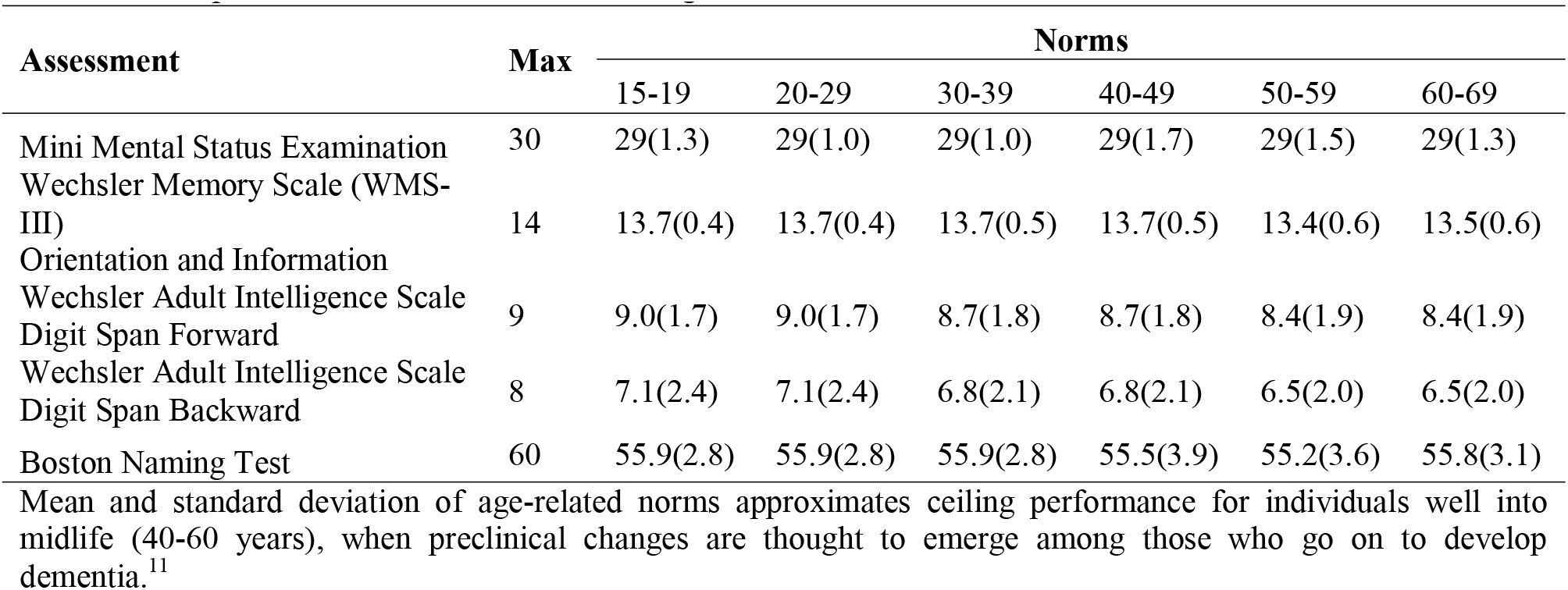
Example normative values of common cognitive tests.

1. What is the relative sensitivity and specificity of a battery of standardized tests frequently used to assess for MCI (comparable to the National Alzheimer’s Coordinating Center Uniform Dataset), as defined using receiver operating characteristic (ROC)-based analysis?
2. What are the optimal “cut point” scores for distinguishing patients’ mildly impaired performance based on these instruments?

Our hypothesis was that assessments designed to have no clear maximum (e.g., phonological fluency) or a very high maximum relative to common performance (e.g., delayed recall) would outperform more commonly utilized, often abbreviated assessments with relatively low maximum possible scores. This hypothesis was informed by our prior analyses of the National Alzheimer’s Coordinating Center frontotemporal lobar degeneration module when used to detect primary progressive aphasia,^9, 10^ in which a similar pattern of results was observed.

## Materials and Methods

We report how we determined our sample size, all data exclusions, all inclusion/exclusion criteria, whether inclusion/exclusion criteria were established prior to data analysis, all manipulations, and all measures in the study.

### Records reviewed

Diagnosis of mild cognitive impairment was determined using a battery of standardized tests (described below), in conjunction with comprehensive neurological evaluation, review of medical history, and consultation with family, if present. The findings of this evaluation were affirmed with magnetic resonance imaging for all patients to rule out the presence of a clear neurological etiology to explain the change observed.

Patients seen in the Johns Hopkins Outpatient Center between under investigation for cognitive impairment were examined retrospectively. Three hundred evaluations of 255 patients seen between December 2018 July 2023 were selected at random. All completed assessments were considered for analysis. No part of the study procedures or analysis plans was pre-registered prior to the research being undertaken. Sixty evaluations were removed due to a known etiology that accounted for cognitive impairments observed in testing: attention deficit disorder (2), cognitive impairment secondary to stroke (26), cerebral amyloid angiopathy (2), anoxic brain injury (4), traumatic brain injury (5), arteriovenous malformations treatments (3), other known conditions resulting in cognitive deficits (11), and other cognitive disorders (7). The remaining patients contributing 240 evaluations are summarized in Table 2. On Fisher-Freeman-Halton Exact Tests, groups were similarly dichotomized in gender distribution and psychiatric comorbidities. While individuals within normal limits were younger than those with clinical diagnoses of either MCI (t(77.7)=3.5, p < 0.001) or dementia (t(116)=2.6, p=0.009), those with clinical profiles did not significantly differ in age.

**Table 2:** Sample characteristics.

|  | **WNL** | **MCI** | **Dementia** |
| --- | --- | --- | --- |
| N individuals | 46 | 96 | 57 |
| N visits | 52 | 122 | 66 |
| Age Mean(SD) | 65(16) | 73(12) | 72(12) |
| Sex (F:M) | 19:27 | 52:44 | 32:25 |
| Psychiatric comorbidities (count) | 15 | 34 | 27 |
| Anxiety | 8 | 13 | 14 |
| Depression | 7 | 19 | 11 |
| Other | 0 | 2 | 2 |

### Assessment battery

A battery of widely used assessments of cognition was employed for each evaluation, the Hopkins Cognitive Battery. The battery contains both simple questions and those designed to probe complex of cognitive demands, even if at the expense of brevity (Table 3). The Mini Mental Status Examination (MMSE) was most commonly used until 2019, when it was replaced by the Wechsler Adult Intelligence Scale--Third Edition (WAIS-III) Wechsler Memory Scale (WMS) Orientation and Information questions.^12^

**Table 3:** Assessment summary.

| **Capacity** | **Hopkins Cognitive Battery** |
| --- | --- |
| Cognitive screening | Mini Mental Status Examination (MMSE)<sup>13</sup><br>WMSIII Orientation & Information questions (WMSIII)<sup>12</sup> |
| Verbal memory | Digits forward and backward<br>Rey Auditory Verbal Learning Test<sup>14</sup> (RVLT) |
| Verbal fluency | 1-minute phonemic fluency (F, A, S)<sup>15</sup><br>Timed written alphabet |
| Naming | 30-item Boston Naming Test (BNT)<sup>16</sup> |
| Writing | Written *Cookie Theft* picture description<sup>17</sup> |
| Visuospatial skills | Trail Making Test (TMT)<br>Rey-Osterrieth Complex Figure Copy<sup>18, 19</sup> |
| Executive Functions | Trail Making Test (TMT)<br>Stroop test<sup>20</sup> |

## Statistical analysis

### Aim 1

The first aim of this work was to examine the relative discriminatory performance of the assessments within the Hopkins Cognitive Battery with regard to sensitivity and specificity across the three contrasts. First, we examined WNL versus MCI+Dementia combined, as determining the presence of clinical change is the most consistent with the scenario when a patient presents in an outpatient clinic under investigation of cognitive change. Next, we examined the two constituent contrasts, WNL versus MCI and WNL versus Dementia, separately. Each visit was treated as an independent, not repeated, portrayal of clinical characteristics. This maximized the overall size of the sampling used for discriminatory analysis, but ignored the potential intra-patient influence; however, 29 patients contributed more than one visit to the analysis.

As in our prior analysis of detection using NACC battery assessments,^9, 10^ calculations were done in R using plotROC 2.2.^21^ to calculate empirical ROC curves and cutpointr for optimizing the cutoff score^22^ on the unsmoothed curve. Receiver operating characteristic (ROC) analysis illustrates the binary diagnostic ability (present or absent) of a given task as the critical threshold score is varied. In this analysis, 0.5 represents chance and 1.0 represents perfect performance. The x-axis is the false positive rate (FPR, also 1-specificity). The y-axis is the true positive rate (TPR, also called sensitivity). Area under the receiver operating characteristic curve (AUC) is a common way to define the overall performance of that measure in performing the diagnostic classification. AUROC 0.7/1 was used to vet tasks most useful to detection, as this is widely considered acceptable performance of a task.^23^ The tasks with acceptable performance detecting MCI versus healthy performance then were entered into a binomial logistic regression. This was done to estimate the overall diagnostic utility of the subset of selected tasks in predicting MCI status. The significance of the model was considered against α = 0.05.

### Aim 2

The second aim of this work was to identify the optimal cut-point scores for detection of MCI based on these instruments. In order to examine Aim 2, “optimal” cut-points were established by maximizing the Youden-Index (sensitivity + specificity – 1)^24^ for any task with a AUC of ≥ 0.7 in the detection of MCI (WNL vs MCI). These values provide insights into scores where specificity and sensitivity interests are balanced, and can be further described in terms of accuracy, sensitivity, and specificity at that score.

### Standard Protocol Approvals, Registrations, and Patient Consents

All work was conducted with the formal approval of the Johns Hopkins University School of Medicine Institutional Review Board (IRB-3; Federal Wide Assurance # FWA00005752-JHUSOM, FWA00006087-JHH & JHHS, and FWA00005719-KKI OHRP IRB Registration #IRB00400111). Participant consent was not required.

## Data Availability

The conditions of our ethics approval do not permit public archiving of study data. Anonymized data not otherwise present in the appendix are available upon request to the authors, subject to review by the Johns Hopkins University School of Medicine Institutional Review Board resulting in a formal data sharing agreement.

## Results

### Aim 1: Receiver Operating Characteristic (ROC) curves

Results of the ROC analysis are presented in Table 4 (Aim 1). While clinical change (either MCI or dementia) was acceptably detected by most administered tasks, this was largely driven by very high AUCs representing the larger magnitude of difference between normal performance (WNL) and performance consistent with a diagnosis of dementia. This is consistent with the fact that these batteries and individual tasks are designed with magnitudes of change consistent with dementia in mind. However, relatively few tasks performed acceptably in the detection of MCI, as defined by the *a priori* cutoff of 0.7 (Table 4, bolded). Notably, those that did perform well tended to have higher total possible scores or not to reflect a score out of a total number possible. This was consistent with our hypothesis regarding the tasks that would be best for identifying MCI.

**Table 4:** Area under the receiver operating characteristic (ROC) curves.

| Task | WNL v Clinical |  | WNL v MCI |  | WNL v Dementia |  |
| --- | --- | --- | --- | --- | --- | --- |
|  | N | AUC | N | AUC | N | AUC |
| MMSE | 61 | 0.77 | 40 | 0.67 | 28 | 0.92 |
| WMSIII orientation | 227 | 0.75 | 165 | 0.67 | 113 | 0.90 |
| Digits forward | 229 | 0.66 | 167 | 0.64 | 113 | 0.70 |
| Digits backward | 227 | 0.76 | 166 | **0.72** | 112 | 0.84 |
| Figure Copy Direct | 160 | 0.75 | 122 | **0.71** | 77 | 0.85 |
| Figure Copy Immediate | 153 | 0.86 | 118 | **0.83** | 73 | 0.94 |
| Figure Copy Delayed | 134 | 0.88 | 107 | **0.86** | 62 | 0.93 |
| RVLT sum | 222 | 0.87 | 166 | **0.84** | 108 | 0.95 |
| RVLT Immediate Recall | 221 | 0.87 | 165 | **0.84** | 108 | 0.94 |
| RVLT Delayed Recall | 212 | 0.89 | 161 | **0.87** | 102 | 0.95 |
| BNT | 221 | 0.76 | 163 | 0.69 | 107 | 0.91 |
| FAS word fluency | 219 | 0.77 | 161 | **0.71** | 109 | 0.89 |
| TMT-A (sec) | 94 | 0.68 | 148 | 0.64 | 93 | 0.76 |
| TMT-B (sec) | 187 | 0.75 | 148 | **0.72** | 87 | 0.84 |
| Stroop Color | 34 | 0.41 | 27 | 0.41 | 11 | 0.63 |
| Stroop Color-Word | 32 | 0.83 | 25 | **0.85** | 11 | 0.79 |
| Alphabet (sec) | 168 | 0.64 | 126 | 0.60 | 80 | 0.73 |
| Alphabet Errors | 171 | 0.62 | 127 | 0.58 | 82 | 0.31 |
| *Cookie Theft* description | 163 | 0.71 | 119 | 0.67 | 79 | 0.78 |

AUC above 0.7 are bolded.

As planned, acceptably performing tasks were entered into a logistic regression to determine how they performed in concert with one another when predicting MCI status versus healthy performance. However, modifications were made based on the limitations of the data as observed *a posteriori*. To reduce the number of dimensions and multicollinearity among predictors, only the best performing score from a given test where multiple scores qualified was entered: digits backward, Rey delayed figure copy, RVLT delayed recall, phonological fluency in FAS, Trail-making test B, and Stroop Color-Words. Second, as very few patients completed the Stroop task, this was removed to maximize the available data with which to calculate the model (22 versus 103).

Overall, the model was significant (χ2(5) = 60.06, p < 0.001), accounting for approximately 61% of the variance in MCI status (Nagelkerke’s R^2^) and classifying 85.4% of cases correctly (Table 5). Holding each of the other tests constant, the odds of performing consistent with MCI decreased by 13% with each additional point on delayed figure copy and by 23% with each additional point on the RVLT delayed recall. No other predictors were independently significant.

**Table 5:**
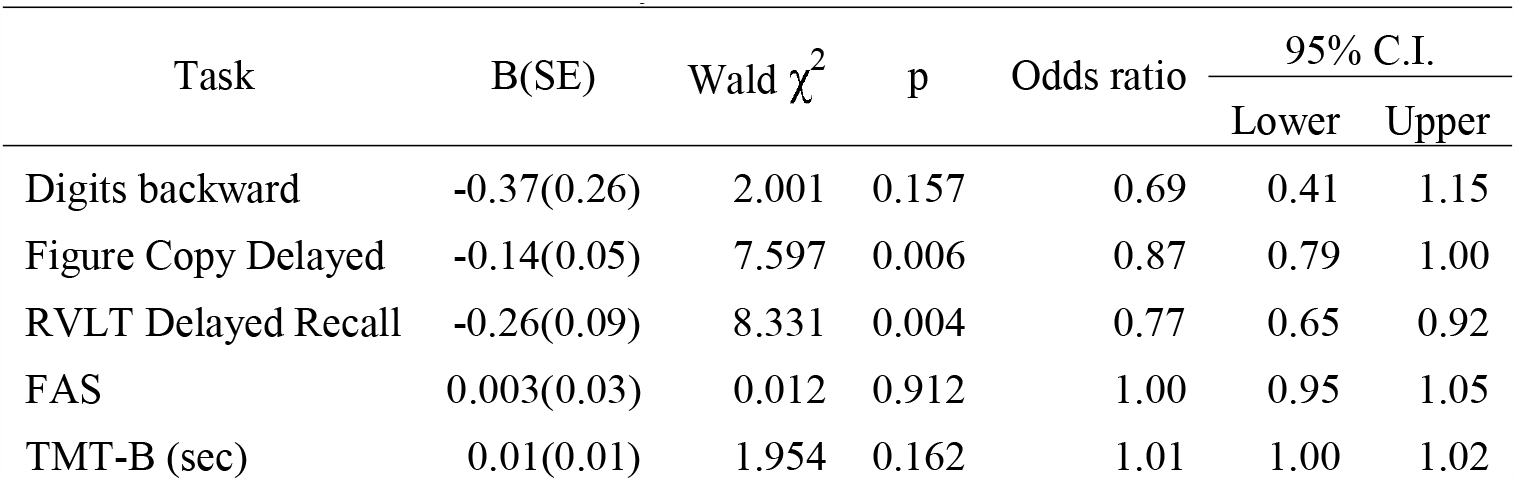
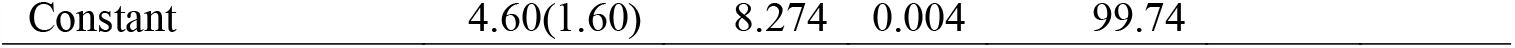
Logistic regression summary.

### Aim 2: Optimal cut-point scores for detecting MCI

Optimal scores were calculated for acceptable tasks based on detection of MCI versus healthy performance. While these calculations are not age-corrected, an important observation is how many fall *within* one standard deviation of normal performance for individuals 60-69 years old (Table 6 †) and all accept the optimal score for Stroop color-word performance fall within one standard deviation of normal performance for individuals 70-79 years old. This highlights the difficulty in identifying MCI on the basis of behavioral testing using common measures for the detection of dementia in older adults.

**Table 6:** Optimal critical scores for identifying MCI versus healthy performance.

| Task | N | Optimal score | Accuracy | Sensitivity | Specificity |
| --- | --- | --- | --- | --- | --- |
| Digits backward | 166 | 4/8 | 0.68 | 0.69 | 0.67 |
| Figure Copy Direct | 122 | 29.5/36† | 0.66 | 0.64 | 0.69 |
| Figure Copy Immediate | 118 | 8/36 | 0.75 | 0.68 | 0.90 |
| Figure Copy Delayed | 107 | 7.5/36† | 0.78 | 0.71 | 0.91 |
| RVLT sum | 166 | 36/75 | 0.77 | 0.79 | 0.73 |
| RVLT Immediate Recall | 165 | 5/15 | 0.77 | 0.75 | 0.81 |
| RVLT Delayed Recall | 161 | 4/15 | 0.76 | 0.73 | 0.84 |
| FAS | 161 | 44† | 0.75 | 0.88 | 0.47 |
| TMT-B (sec) | 148 | 32.6† | 0.74 | 0.84 | 0.52 |
| Stroop Color-Word | 25 | 53 | 0.76 | 0.71 | 1.00 |

† *Within* one standard deviation of normal performance for individuals 60-69 years old.

## Discussion

When patients present for an outpatient neurological evaluation due to concerns regarding perceived cognitive change, they most commonly receive assessments designed with the detection of dementia in mind. However, based on our sample, nearly half (48%) of those who presented showed abnormal performance in at least one domain and otherwise met criteria for MCI, rather than dementia. To better understand task performance among this etiologically diverse and poorly understood population, we pursued two aims using a large retrospective sample of outpatient data. First, we examined the acceptability of individual commonly administered tasks in a receiver operating characteristic (ROC) analysis. Our hypothesis was that the tasks that performed best would be those in which the most common scores were well below ceiling performance (and well above floor) either due to high numbers of points possible or no maximum score (i.e., counts). Next, we used the acceptability threshold to calculate a binomial logistic regression determining how tasks worked together to detect MCI. Finally, we calculated optimal scores for distinguishing between performance of healthy individuals and performance of those with MCI. To assist in the interpretation of these values, we briefly contrasted them with available norms.

Based on our ROC analysis, the Hopkins Cognitive Battery tasks performed well in the detection of clinical change generally. However, this was driven by nearly perfect detection of dementia, rather than the milder deficits associated with MCI. Our hypothesis regarding which tasks would perform best in detecting MCI was supported; high performing tasks tended to be those like the Rey Complex Figure Copy and Rey Auditory Verbal Learning Test, which both are quite difficult, and thus, normal performance is well below ceiling performance. The number of words produced beginning with F, A, and S and amount of time needed to complete the Trail-making Test Part B time also performed well in detection of MCI. While Stroop Color-Word naming (that is, naming the color the word, such as BLUE, is printed in), also performed well, relatively few instances of performance were available for analysis of this assessment. Thus, it should be interpreted thoughtfully.

Particularly unfortunate was that the widely used cognitive screening tool, the MMSE, did not perform well in the detection of MCI. Perhaps less surprisingly, the Wechsler Memory Scale (WMS) Orientation and Information questions did not perform well either. This is an important observation for its practical clinical significance. If a patient comes in describing perceived cognitive change, and their performance on these abbreviated measures is within normal limits for their age, it could easily lead to missed identification of true MCI by well-meaning generalists, and those with MCI who do progress to dementia would have missed out on having an earlier understanding of their risk. A more conservative approach that is supported by the strong performance of tests more commonly used by behavioral neurologists and neuropsychologist would be to provide a referral for full cognitive evaluation for these patients. In that setting, an extended evaluation utilizing multiple high performing assessments is possible and may provide corroborative evidence of the patients’ experience or, conversely, provide a comprehensive personal baseline for a healthy aging individual that can be referenced in years to come.

Examining optimal cutoff scores establishes a useful guideline for critically examining a patient’s profile assessment by assessment. However, as previously noted, many of these scores would be considered typical of healthy individuals even as young as 60. This finding highlights two important points regarding interpretation. First, among those tested, no one task in isolation is sufficient to detect MCI, as expected because there are distinct types of MCI (amnestic, non-amnestic, single domain, multiple domain). Furthermore, behavioral performance is not the only relevant consideration in differential diagnosis. A more sensitive and specific approach to detection likely requires task performance to be considered in conjunction with a review of medical history, consultation with family, and, if possible, neuroimaging. These findings also reinforce that the currently available assessments and subsequent norms, most based only on age, have limited utility in detecting very mild impairments. As we become increasingly capable of treating the underlying etiologies that lead to cognitive impairments, the number of people whose impairments are mild and who are less frequently fully disabled by such impairments likely will increase. The lack of assessments in widespread use that can detect such impairments and the poverty of individualized norms available for well-established tests is an important area of future innovation.

One limitation of this work was that the number of instances of certain tasks available for analysis was quite low (e.g., Stroop). These tasks are administered relatively infrequently relative to the remainder of the battery. The Stroop task was not administered systematically during the coronavirus pandemic, when assessments were conducted almost exclusively using telemedicine and video conferencing. Even in person, the Stroop is not appropriate for many individuals, including those with color blindness or presbyopia. Future work should expand upon the analyses of these instruments in a larger sample.

Overall, these findings represent a positive impression of frequently used tests in accomplishing its key purpose: detecting dementia. However, they also highlight the relative weakness of our most administered assessments when used to build a comprehensive profile of a very large portion of outpatients presenting at clinic, those whose deficits are more subtle. It is our hope that this work will galvanize future clinicians and clinical researchers to develop a better understanding of MCI and the heterogeneous individuals who experience subtle cognitive changes beyond what is expected for their age.

## Data Availability

The conditions of our ethics approval do not permit public archiving of study data. Anonymized data are available upon request to the authors, subject to review by the Johns Hopkins University School of Medicine Institutional Review Board resulting in a formal data sharing agreement.

## References

1. Albert MS, DeKosky ST, Dickson D, et al. The diagnosis of mild cognitive impairment due to Alzheimer’s disease: recommendations from the National Institute on Aging-Alzheimer’s Association workgroups on diagnostic guidelines for Alzheimer’s disease. Alzheimers Dement. May 2011;7(3):270–9. doi:10.1016/j.jalz.2011.03.008

2. Lezak MD. Neuropsychological assessment. Oxford University Press, USA; 2004.

3. Busch RM, Chapin JS. Review of normative data for common screening measures used to evaluate cognitive functioning in elderly individuals. The Clinical Neuropsychologist. 2008;22(4):620–650.

4. Busch RM, Chelune GJ, Suchy Y. Using norms in neuropsychological assessment of the elderly. Geriatric neuropsychology: Assessment and intervention. 2006:133–157.

5. Manly JJ, Echemendia RJ. Race-specific norms: Using the model of hypertension to understand issues of race, culture, and education in neuropsychology. Arch Clin Neuropsychol. 2007;22(3):319–325.

6. Park C, Jang J-W, Joo G, et al. Predicting progression to dementia with “comprehensive visual rating scale” and machine learning algorithms. Original Research. Front Neurol. 2022-August-22 2022;13doi:10.3389/fneur.2022.906257

7. Chen J, Chen G, Shu H, et al. Predicting progression from mild cognitive impairment to Alzheimer’s disease on an individual subject basis by applying the CARE index across different independent cohorts. Aging (Albany N Y). 2019;11(8):2185.

8. Pereira T, Lemos L, Cardoso S, et al. Predicting progression of mild cognitive impairment to dementia using neuropsychological data: a supervised learning approach using time windows. BMC Medical Informatics and Decision Making. 2017/07/19 2017;17(1):110. doi:10.1186/s12911-017-0497-2

9. Stockbridge MD, Tippett DC, Breining BL, Vitti E, Hillis AE. Task performance to discriminate among variants of primary progressive aphasia. Cortex. 2021/12/01/ 2021;145:201–211. doi:10.1016/j.cortex.2021.09.015

10. Stockbridge MD, Tippett DC, Breining BL, Hillis AE. When words first fail: Predicting the emergence of primary progressive aphasia variants from unclassifiable anomic performance in early disease. Aphasiology. 2023;37(8):1173–1185. doi:10.1080/02687038.2022.2084706

11. Hou Y, Dan X, Babbar M, et al. Ageing as a risk factor for neurodegenerative disease. Nature Reviews Neurology. 2019;15(10):565–581.

12. Wechsler D. Wechsler Adult Intelligence Scale--Third Edition (WAIS-III). Psychological Corporation; 1997.

13. Folstein MF, Folstein SE, McHugh PR. “Mini-mental state”: a practical method for grading the cognitive state of patients for the clinician. J Psychiatr Res. 1975;12(3):189–198.

14. Schmidt M. Rey auditory verbal learning test: A handbook. vol 17. Western Psychological Services Los Angeles, CA; 1996.

15. Lezak M. Neuropsychological Assessment. 3rd ed. Oxford University Press; 1995.

16. Williams BW, Mack W, Henderson VW. Boston naming test in Alzheimer’s disease. Neuropsychologia. 1989;27(8):1073–1079. doi:10.1016/0028-3932(89)90186-3

17. Goodglass H, Kaplan E, Weintraub S. BDAE: The Boston diagnostic aphasia examination. Lippincott Williams & Wilkins Philadelphia, PA; 2001.

18. Rey A. L’examen psychologique dans les cas d’encephalopathie traumatique. Arch Psychol (Geneve). 1941;28:286–340.

19. Osterrieth PA. Le test de copie d’une figure complexe; contribution a l’etude de la perception et de la memoire. Arch Psychol (Geneve). 1944;

20. Stroop JR. Studies of interference in serial verbal reactions. J Exp Psychol. 1935;18(6):643.

21. Sachs MC. plotROC: a tool for plotting ROC curves. Journal of statistical software. 2017;79

22. Thiele C, Hirschfeld G. Cutpointr: Improved estimation and validation of optimal cutpoints in R. arXiv preprint arXiv:200209209. 2020;

23. Hosmer DW, Lemeshow S, Sturdivant RX. Applied logistic regression. vol 398. John Wiley & Sons; 2013.

24. Perkins NJ, Schisterman EF. The Youden Index and the optimal cutLJpoint corrected for measurement error. Biometrical Journal: Journal of Mathematical Methods in Biosciences. 2005;47(4):428–441.

